# mRNA COVID-19 vaccine elicits potent adaptive immune response without the persistent inflammation seen in SARS-CoV-2 infection

**DOI:** 10.1101/2021.04.20.21255677

**Authors:** Ellie N. Ivanova, Jasmine Shwetar, Joseph C. Devlin, Terkild B. Buus, Sophie Gray-Gaillard, Akiko Koide, Amber Cornelius, Marie I. Samanovic, Alberto Herrera, Eleni P. Mimitou, Chenzhen Zhang, Trishala Karmacharya, Ludovic Desvignes, Niels Ødum, Peter Smibert, Robert J. Ulrich, Mark J. Mulligan, Shohei Koide, Kelly V. Ruggles, Ramin S. Herati, Sergei B. Koralov

## Abstract

SARS-CoV-2 infection and vaccination elicit potent immune responses. Our study presents a comprehensive multimodal single-cell dataset of peripheral blood of patients with acute COVID-19 and of healthy volunteers before and after receiving the SARS-CoV-2 mRNA vaccine and booster. We compared host immune responses to the virus and vaccine using transcriptional profiling, coupled with B/T cell receptor repertoire reconstruction. COVID-19 patients displayed an enhanced interferon signature and cytotoxic gene upregulation, absent in vaccine recipients. These findings were validated in an independent dataset. Analysis of B and T cell repertoires revealed that, while the majority of clonal lymphocytes in COVID-19 patients were effector cells, clonal expansion was more evident among circulating memory cells in vaccine recipients. Furthermore, while clonal αβ T cell responses were observed in both COVID-19 patients and vaccine recipients, dramatic expansion of clonal γδT cells was found only in infected individuals. Our dataset enables comparative analyses of immune responses to infection versus vaccination, including clonal B and T cell responses. Integrating our data with publicly available datasets allowed us to validate our findings in larger cohorts. To our knowledge, this is the first dataset to include comprehensive profiling of longitudinal samples from healthy volunteers pre/post SARS-CoV-2 vaccine and booster.

## Introduction

In 2019, the novel coronavirus SARS-CoV-2 emerged, and the resulting pandemic has had unprecedented impact on the heath, economy, and social fabric of the global community. The clinical presentation of coronavirus disease 2019 (COVID-19), the disease caused by SARS-CoV-2, has been highly heterogeneous, with manifestations ranging from asymptomatic or mild illness to acute respiratory distress syndrome (ARDS), multiorgan failure, and death.

To date, a number of comprehensive studies have described immune responses to SARS-CoV-2 infection (Carsetti et al., 2020; Mathew et al., 2020; Mohammed et al., 2022; Stephenson et al., 2021; Wilk et al., 2020). These research efforts identified lymphopenia with concomitant innate cell expansion, while specific alterations in a number of immune subsets, including activated CD8 T cells, plasma cells, monocytes, and NK cells, are thought to shape patients’ clinical outcomes. Immune responses in individuals who survive COVID-19 eventually return to baseline, with establishment of memory T and B cell responses (Kared et al., 2021; Laidlaw and Ellebedy, 2022; Moss, 2022; Pape et al., 2021; Turner et al., 2021a; Turner et al., 2021b; Zhang et al., 2020), and corresponding development of a neutralizing antibody repertoire (Jeewandara et al., 2021; Wang et al., 2020).

Infection with SARS-CoV-2 and vaccination against the virus have both been shown to stimulate immune responses and to protect against subsequent infection (Chodick et al., 2021; Dan et al., 2021; Lederer et al., 2020; Moss, 2022; Thompson et al., 2021; Turner et al., 2021b). Both infection and vaccination generate protective anti-S memory immune responses (Chodick et al., 2021; Dan et al., 2021; Thompson et al., 2021). A detailed comparison of immune responses to the virus versus vaccination and subsequent boosters provides a unique opportunity to contrast immune reactions linked to infection and immunization with well-defined antigens.

In our study, we took advantage of exCITE-seq (<u>ex</u>panded <u>C</u>ellular Indexing of <u>T</u>ranscriptomes and <u>E</u>pitopes by <u>seq</u>uencing), a multimodal, single-cell sequencing technique, to simultaneously characterize the surface phenotype and transcriptome of immune cells (Mimitou et al., 2019; Stoeckius et al., 2017). This platform also enables reconstruction of B cell and T cell antigen receptor rearrangement of individual lymphocytes. We used the exCITE-seq platform to characterize cellular and transcriptional responses to SARS-CoV-2 infection and vaccination in peripheral immune cells to better understand the host response to the pathogen and to immunization against defined viral antigens. We also quantified virus-specific antibody titers in the serum of COVID-19 patients and vaccine recipients using a recently developed multiplex bead-binding assay (Hattori et al., 2021) and antibody ELISA (Samanovic et al., 2022).

Our multimodal analysis revealed dramatic alterations in the frequencies and transcriptional programs of several immune subsets in response to infection and highlighted differences in the breadth of immune responses observed upon infection and vaccination. In COVID-19 patients, transcriptional profiles of many immune cell populations were characterized by augmented IFN signaling, upregulation of genes associated with cytotoxicity, and changes in metabolic pathways. These transcriptional changes were also readily observed in an independent scRNA-seq dataset of 38 acute and 38 convalescent patients, with the cytotoxic signature persisting in circulating cells of convalescent patients (Stephenson et al., 2021). Analysis of peripheral immune cells following vaccination with the BNT162b2 mRNA vaccine also revealed alterations of transcriptional programs of several immune cell populations consistent with immune activation, but the highly augmented interferon (IFN) signaling and cytotoxic signature observed in COVID-19 patients were largely absent. We observed robust antibody response in both COVID-19 patients and immunized individuals, with vaccination inducing a remarkably consistent IgG response to S protein. Interestingly, B cell and T cell clonal responses differed dramatically between infected and vaccinated individuals, suggesting that infection-driven inflammation influences the trajectory of the adaptive immune response; this may have important implications for our understanding of the durability of protective immune responses.

## Results

### Overview of immune responses to COVID-19 infection and immunization

To improve our understanding of immune responses to SARS-CoV-2 antigens in different inflammatory contexts, we profiled circulating immune cells from five adults with acute COVID-19 and nine healthy adults, seven of whom received the BNT162b2 vaccine, and two who were SARS-CoV-2 naïve. For three of the vaccine recipients, samples were also collected before and after receiving a booster. Samples were taken at multiple time points, resulting in a total of 42 post-vaccination and 9 post-infection samples (Schematic 1). COVID-19 samples were collected during the acute phase of infection. For four of the five COVID-19 patients, we obtained longitudinal samples. COVID-19 sample time points were recorded as days post-onset of symptoms, and clinical metadata were evaluated for clinical severity based on the WHO clinical progression scale, in which 1-3 represents mild, 4-5 moderate, and 6-9 severe disease (WHO Working Group on the Clinical Characterisation and Management of COVID-19 Infection, 2020). All subjects in the vaccinated group received two doses approximately three weeks apart, in accordance with its FDA Emergency Use Authorization. For vaccine responses, samples were collected at baseline, and then at approximately 1, 3, and 4 weeks after the first vaccine dose. For three of the vaccine recipients, we collected samples at additional time points: at 5 weeks post-vaccine as well as pre-booster, and 1 and 4 weeks post-booster. For all participants, demographic characteristics, clinical features, and outcomes are listed in Supplemental Table 1. The age distribution of study participants was similar in vaccine and COVID-19 groups (Supplemental Fig. 1).

**Schematic 1.**
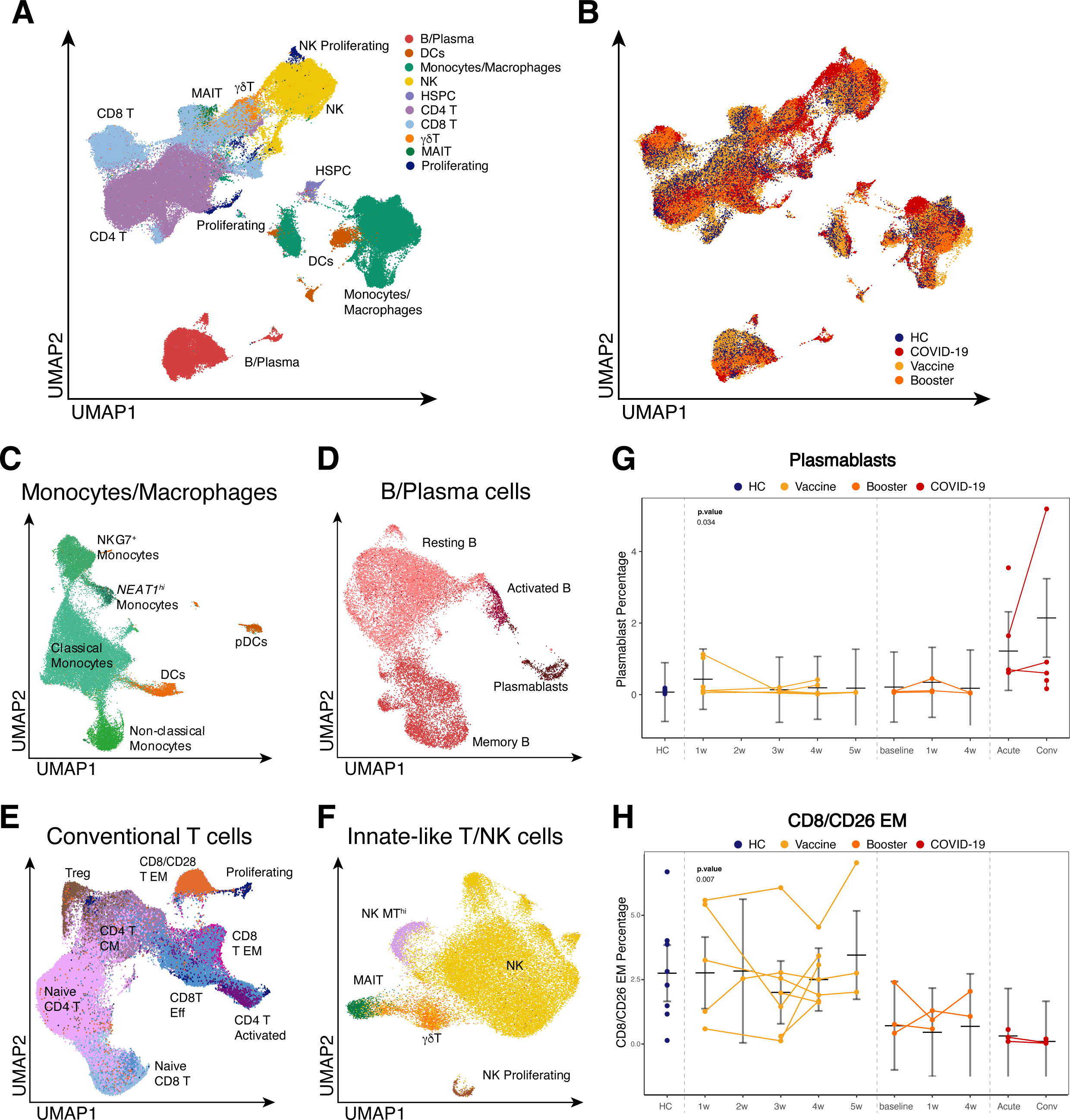

To assess the impact of SARS-CoV-2 infection and vaccination on each individual’s global immune landscape, we used a multimodal exCITE-seq approach (Mimitou et al., 2019) to identify discrete clusters based on the transcriptional profile and surface epitopes of circulating cells. To do this, peripheral blood mononuclear cells (PBMCs) were multiplexed and processed using 5’ droplet-based scRNA-seq technology (10x Genomics). Surface marker phenotypes were detected using an optimized 60-antibody CITE-seq panel (Buus et al., 2020), generating matching transcriptional and surface protein data. In addition, for each sample, we sequenced single-cell T cell receptors (TCR) αβ and γδ, as well as B cell receptors (BCRs), to evaluate antigen receptor repertoires. Samples from healthy volunteers prior to vaccination were grouped together with samples from unvaccinated COVID-19-naïve healthy donors as healthy controls (HC).

In total, we obtained 195,634 PBMCs from 51 individual samples, with an average of 3,600 cells/sample. Among these, 36,906 cells (∼19%) were from COVID-19 patients; 37,680 cells (∼19%) were from HCs and pre-vaccine samples; 90,753 cells (∼46%) were from post-vaccine samples; and 30,295 cells (∼15%) came from booster samples. All high-quality single cells were integrated across the RNA, antibody-derived tags (ADTs), TCR and BCR modalities for all subsequent analyses. Dimension reduction was performed using the combined RNA and ADT modalities to generate a uniform manifold approximation and projection (UMAP) (McInnes and Healy, 2018) representation of all 195,634 cells from HC, and from immunized volunteer and COVID-19 patient samples (Fig. 1A, B). Using a combination of Louvain-based clustering (Blondel et al., 2008), SingleR (Aran et al., 2019) reference-based annotation, and literature markers, we identified 10 major lineages (Fig. 1A, B) and 24 individual subpopulations of myeloid cells, B cells, conventional and innate-like T cells, and NK cells (Fig. 1C-F). Gene expression and canonical ADT markers further confirmed these lineages and sub-populations (Supplemental Fig. 2A,B, Supplemental Table 2).

**Figure 1.**
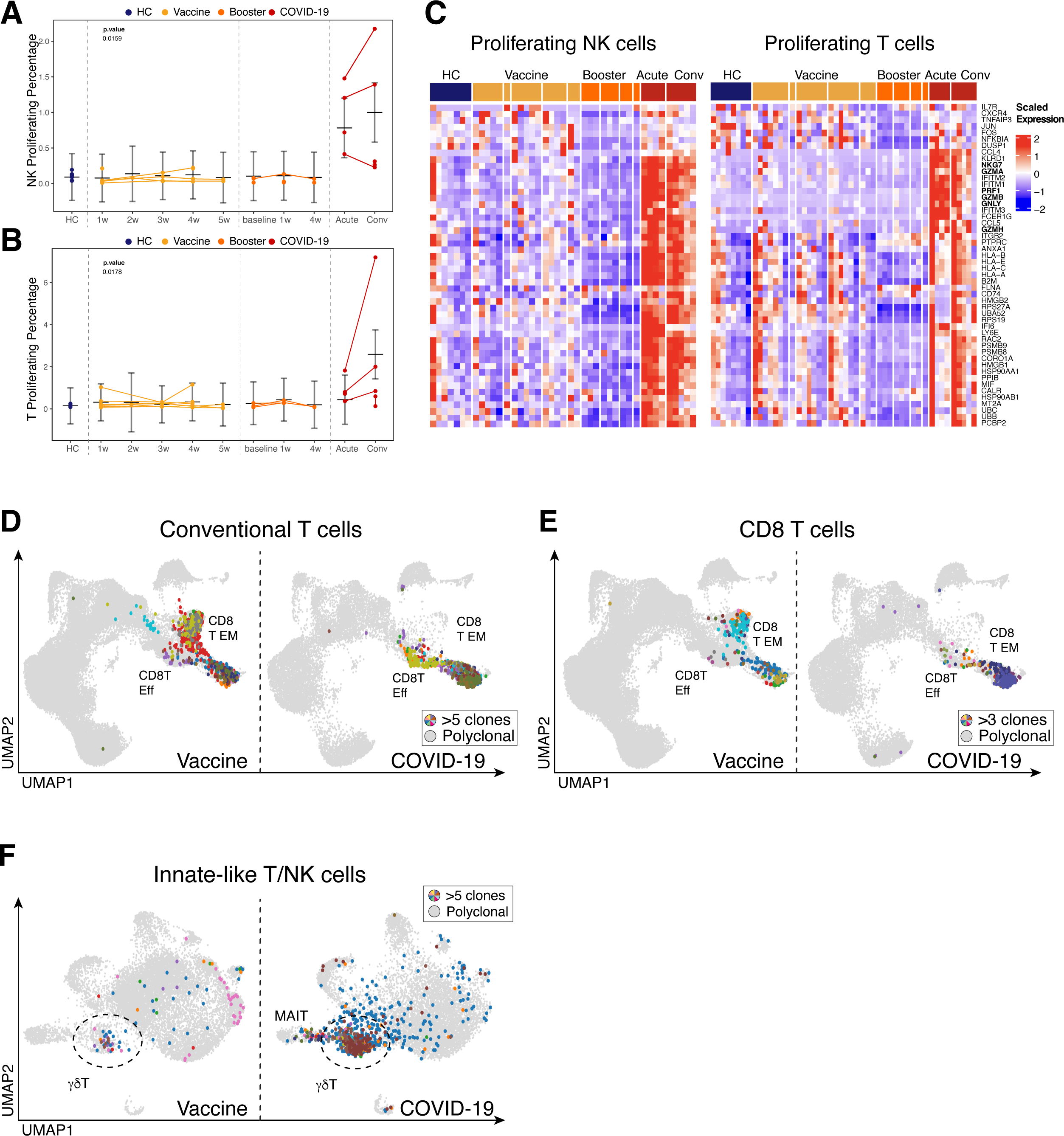

A number of studies have demonstrated a highly heterogenous anti-viral inflammatory responses in COVID-19 patients, likely due to variability of disease severity, stage of disease, and diversity of preexisting conditions (Mohammed et al., 2022; Moss, 2022; Stephenson et al., 2021; Wilk et al., 2020; Zhang et al., 2020). Similarly, our analysis revealed striking differences in the frequency of key immune cell populations between COVID-19 patients and healthy volunteers prior to and following vaccination (Fig. 1G,H).

### Dramatic difference in maturation of B cell responses triggered by SARS-CoV-2 infection and vaccination

Current COVID-19 vaccine efforts have focused on the generation of humoral immune responses against SARS-CoV-2, which has been demonstrated to be a correlate of protection against infection (Finch et al., 2022; Harvey et al., 2020). To better understand the humoral responses following infection and vaccination, we examined B cell responses in the CITE-seq dataset. Single-cell analysis identified four distinct B cell populations based on gene expression and surface epitopes (Fig. 1D). Relative to healthy volunteers, we observed striking expansion of circulating plasmablasts in COVID-19 patients at acute time points (Fig. 1G). In contrast, we observed no apparent expansion of plasmablasts in circulation following vaccination, despite a successful humoral response in all subjects (Supplemental Fig. 3).

As plasmablasts are likely recent emigrants from lymphoid tissue, we hypothesized that they may carry a transcriptional imprint of the inflammatory milieu in tissue. We performed Gene Set Variation Analysis (GSVA) of plasmablasts from COVID-19 patients and healthy volunteers to find out whether signaling pathways were similarly expressed in both cohorts (Fig. 2A,B, Supplemental Fig. 4). This analysis revealed that, relative to plasmablasts in healthy volunteers across all time points, plasmablasts from COVID-19 patients across all time points were highly enriched for genes involved in oxidative phosphorylation, type I and type II IFN responses (IFN-I, IFN-II), fatty acid metabolism, and mTORC1 signaling. The extent of upregulation of IFN response genes in COVID-19 patients correlated with severity of disease, as judged by fraction of inspired oxygen (Fig. 2C). The elevated IFN-I signature is evident in our dataset and was validated using 76 COVID-19 patient samples stratified by disease severity in a large publicly available dataset (Fig. 2B, Supplemental Fig. 5) (Stephenson et al., 2021). Plasmablasts from both COVID-19 patients and healthy volunteers after vaccination had elevated transcription of genes linked to IL-6 receptor signaling (JAK/STAT) and PI3K/AKT signaling pathways (Fig. 2A, Supplemental Fig. 5), two pathways associated with promoting plasmablast differentiation (Fruman and Limon, 2012; Hirano et al., 2000). In contrast to the transcriptional profile of plasmablasts from vaccinated individuals, which resembled that of healthy controls, the increased IFN signaling observed in plasmablasts from acute COVID-19 samples likely reflects the profound inflammation in infected patients. These transcriptional changes in response to IFN and other pro-inflammatory cytokines are likely to have broader implications for B cell differentiation and persistence.

**Figure 2.**
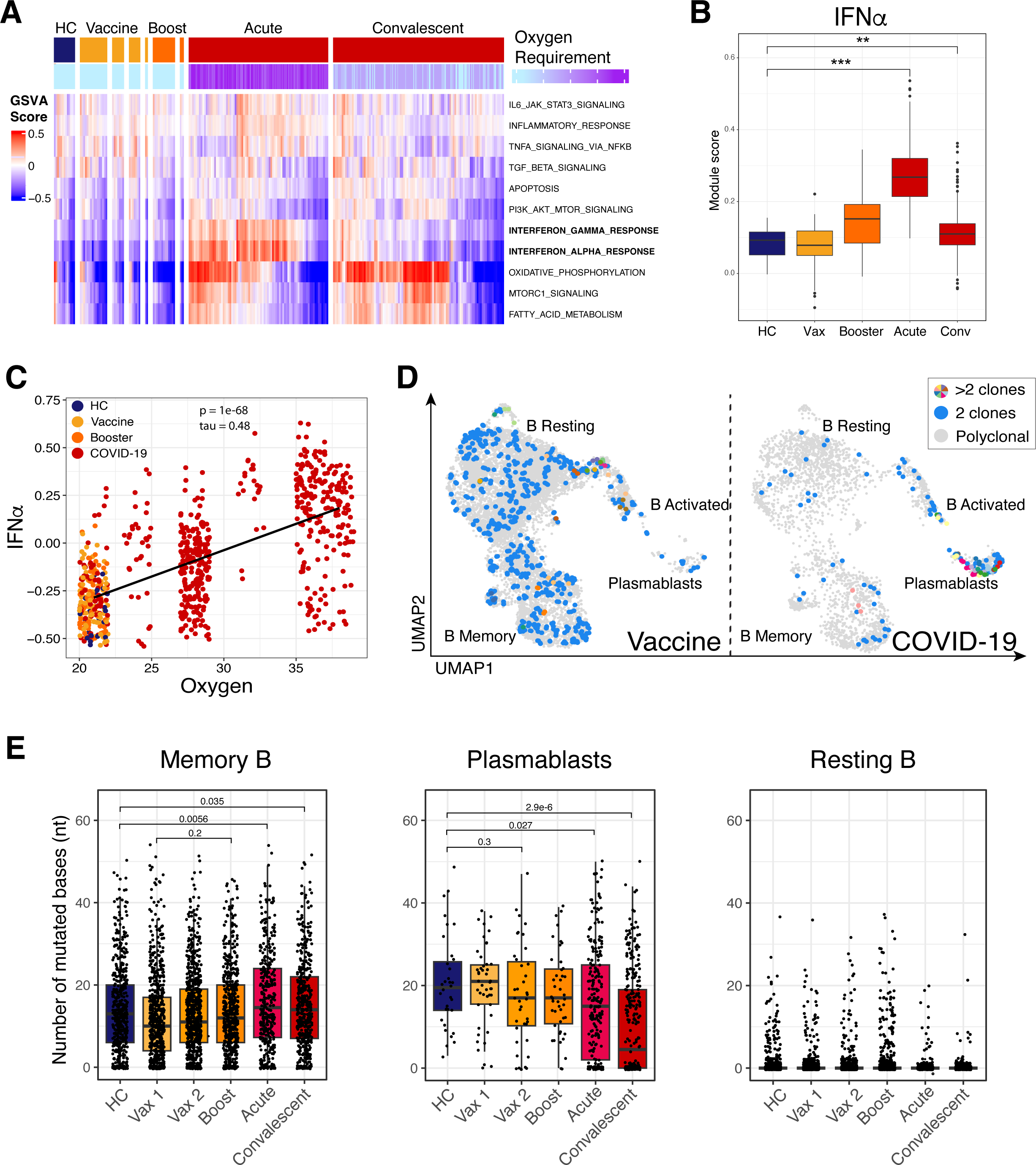

We next evaluated the B cell repertoire across these cohorts. Expansion of B cell clones, as well as convergent antibody repertoires, have been reported for a number of viral infections, including SARS-CoV-2 (Jackson et al., 2014; Nielsen et al., 2020; Robbiani et al., 2020). Analysis of the B cell receptor (BCR) repertoire revealed that a majority of clonal B-lineage cells were captured in the plasmablast cluster in COVID-19 patients. In vaccinated individuals, clonally expanded cells were primarily in resting and memory compartments (Fig. 2D). This observation suggests that the IFN response associated with SARS-CoV-2 infection may promote rapid plasma cell differentiation in COVID-19 patients, while the BNT162b2 mRNA vaccine appears to favor clonal expansion of memory B cells.

To evaluate the extent of somatic hypermutation (SHM), we performed IgBlast-based (Ye et al., 2013) alignment of the V(D)J repertoire for the sequenced samples and then evaluated single base-pair mismatches within the V_H_ alignments for the previously defined B cell clusters. Increased SHM was apparent in memory B cells from all samples, with frequency of mutations notably higher when compared to resting B cells (Fig. 2E). Although few plasmablasts were captured in PBMCs from vaccinated individuals, the rate of SHM in plasmablasts in these samples was comparable to that observed from COVID-19 patients at the peak of disease. Frequency of SHMs was significantly reduced in plasmablasts from convalescent patients’ COVID-19 samples compared to peak disease (Fig. 2E), possibly as a consequence of long-lived plasma cells’ migrating out of circulation (Halliley et al., 2010). It is also possible that some of the antibody-producing plasma cells are either retained at the site of infection or in the draining lymph nodes (MacLean et al., 2022). The observation that Ig rearrangements in plasma cells from healthy individuals carry substantial somatic hyper mutation (SHM) burden is likely a consequence of affinity maturation in response to ongoing immune surveillance, as resting B cells from all individuals had very low background of mutations in their repertoire. There was an increase in SHM within the pool of memory cells one week after a person received the second dose of the vaccine compared to SHM one week after receiving the first dose (Fig. 2E). Despite the inherent limitation of surveying only circulating memory B cells and plasma cells, these analyses highlight ongoing affinity maturation of B cell responses in both COVID-19 patients and vaccinated individuals.

### NK and clonal T cell responses differ in infection and vaccination

Cell-mediated immune responses are carried out by NK cells, CD4, CD8 T cells, and unconventional T lymphocytes like gamma delta (γδ) T cells. Consistent with prior reports, we observed an increase in the frequency of proliferating T cells and NK cells in COVID-19 patients compared to healthy volunteers and vaccinated individuals (Fig. 3A,B) (Di Vito et al., 2022; Hanna et al., 2020). In COVID-19 patients, we measured a dramatically elevated cytotoxic signature in NK cells, CD4 and CD8 T cells, and γ8 T cells (Fig. 3C, Supplemental Fig. 6A). Both CD8 effector T cells and NK cells in COVID-19 patients showed significantly elevated expression of genes associated with cytotoxicity, such as *GZMA*, *GZMB*, *GZMH*, *GNLY*, *NKG7*, and *PRF1* (Figs. 3C, Supplemental Fig. 6A). Because the number of COVID-19 patients in our study was limited, we also verified the elevation of cytotoxic gene signatures in a published dataset of PBMC biospecimens from COVID-19 patients (Supplemental Fig. 6B) (Stephenson et al., 2021). This finding is consistent with previously published results that described a dysregulation of immune responses in COVID-19 (Georg et al., 2022; Yu et al., 2020). Strikingly, while clonal expansion was evident only in CD8 effector T cells of COVID-19 patients, the BNT162b2 vaccine elicited robust clonal responses in both CD8 effector T cells and CD8 T_EM_ cells, suggesting that the vaccine may elicit a more potent memory CD8 response than does infection (Fig. 3D).

**Figure 3.**
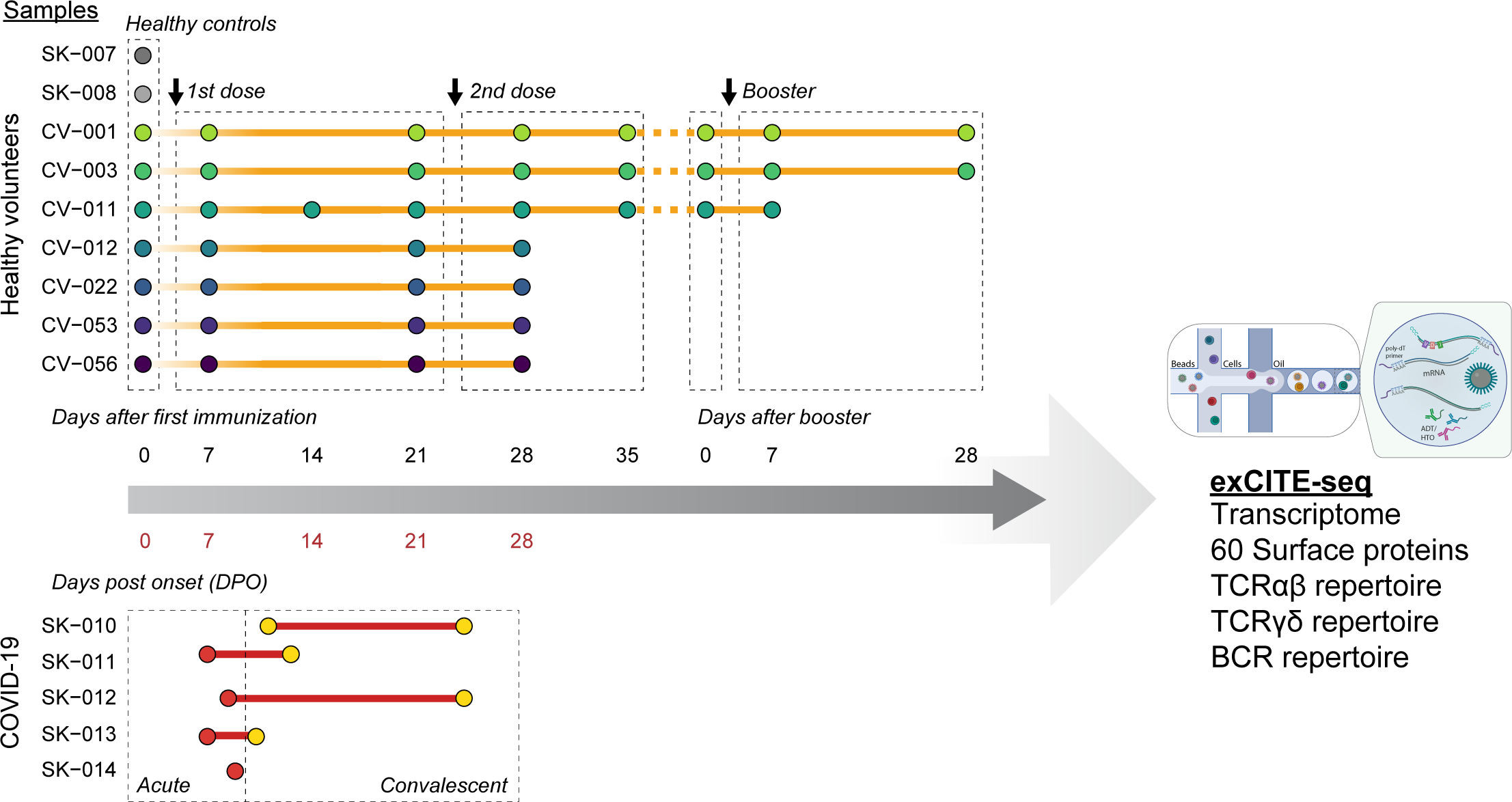

To evaluate whether clonal CD8 cells are SARS-CoV-2 reactive, we mapped CDR3s from a large-scale database of SARS-CoV-2-specific TCRs (Nolan et al., 2020). We found that many clonally expanded CD8 T cells within the CD8 effector T cell and CD8 T_EM_ clusters have CDR3s that perfect match reported SARS-CoV-2-specific TCR sequences (Fig. 3E, Supplemental Fig. 7).

CD4 T helper cells orchestrate much of the adaptive anti-viral immune response. Strikingly, we observed that the majority of activated CD4 T cells from COVID-19 patients expressed genes associated with cytotoxic effector function, such as *GZMH*, *GZMA*, and *PRF1* (Supplemental Fig. 6A,C). Cytotoxic CD4 T cells have been previously observed in COVID-19 and other viral infections (Juno et al., 2017; Kaneko et al., 2021; Meckiff et al., 2020) and have also been noted in patients with autoimmune diseases (Takeuchi and Saito, 2017). Clonally expanded activated CD4 T cells were observed in COVID-19 patients and vaccinated individuals (Fig. 3D), but only COVID-19 patients were characterized by elevated expression of cytotoxic genes (Supplemental Fig. 6A,C). Whether these cells contribute to virus clearance or to inflammation-associated pathology in COVID-19 is unclear.

To evaluate whether clonally expanded T cells in vaccinated individuals and COVID-19 patients in our single-cell sequencing dataset were reactive to SARS-CoV-2, we mapped CDR3s identified via scRNA-seq following *in vitro* activation with recombinant SARS-CoV-2 spike proteins (Gray-Gaillard et al., 2022). A notable number of clonally expanded T cells identified in COVID-19 patients and immunized individuals had matching CDR3s with *in vitro* activated spike-specific T lymphocytes (Supplemental Fig. 8).

Our single-cell analysis also revealed that non-conventional lymphocytes were also engaged by both the viral infection in COVID-19 patients and the mRNA vaccine. Γδ T cells are a subset of unconventional, non-MHC-restricted innate-like T cells with cytotoxic effector functions and the ability to regulate other immune cells (Lawand et al., 2017; Sabbaghi et al., 2020). Transcriptional analysis revealed dramatic upregulation of genes associated with cytotoxic effector functions in the γδ T cells from COVID-19 patients, a feature not observed in γδ T cells from HCs or from vaccinated individuals (Supplemental Fig. 6). Repertoire analysis of γδ T cells revealed oligoclonal expansion in a majority of COVID-19 patients and a moderate dynamic response in vaccinated individuals (Fig. 3F, Supplemental Fig. 9). Overall, our single-cell analysis of lymphocyte responses demonstrated a persistent upregulation of genes associated with cytotoxic effector functions among NK cells, CD4 T, CD8 T, and γδ T cells in COVID-19 patients. The increased cytotoxicity of these cells likely contributes both to pathogen clearance and to immune-mediated pathology.

## Discussion

In this study, we performed a multimodal analysis of samples from COVID-19 patients and from healthy volunteers before and after they received the SARS-CoV-2 BNT162b2 mRNA vaccine and booster. While both infection and immunization elicited robust humoral responses, our analysis revealed dramatic differences in cell composition and transcriptional profiles of circulating immune cells in response to the two different immune challenges.

Type I IFN mediates antiviral immunity, drives expression of a number of genes involved in viral clearance, and plays a critical role in initiating innate and adaptive immune responses during a viral infection (Wu and Metcalf, 2020). IFN signaling induced by viral infection orchestrates antigen presentation, cellular trafficking, and terminal differentiation of lymphocytes (Adamo et al., 2021; Gessani et al., 2014). However, prolonged IFN-I signaling also promotes immunopathology through induction of aberrant inflammatory responses during acute viral infection and can lead to immune dysfunction (McNab et al., 2015). Although the role of IFN-I signaling in COVID-19 awaits full elucidation, recent studies show that systemic production of type I IFN is negatively correlated with disease severity (Hadjadj et al., 2020; Liu et al., 2021), while excessive local production exacerbates lung tissue damage and correlates with increased morbidity and mortality (Broggi et al., 2020). Furthermore, individuals who died of severe COVID-19, which is associated with very high IFN-I, had poor germinal center (GC) responses (Kaneko et al., 2020). While SARS-CoV-2 infection led to a discernible IFN response, we did not find evidence of IFN induction by the BNT162b2 vaccine. This observation suggests that robust affinity maturation in response to viral antigens can occur in the absence of high levels of systemic IFN signaling. It is possible that IFN induction by the vaccine is transient and occurs early and therefore was not captured by our sample collection schedule (Bergamaschi et al., 2021; Li et al., 2022). In addition, current mRNA vaccines incorporate chemically-modified nucleosides to prevent activation of TLR7 and other innate sensors, reducing IFN-I production (Granados-Riveron and Aquino-Jarquin, 2021). Our study lacks the timepoints to evaluate IFN-I signaling immediately after vaccination, but analysis of our exCITE-seq data and of previously published single-cell dataset suggest that infection with SARS-CoV-2 results in profound upregulation of type I IFN signaling.

COVID-19 patients had a striking expansion of antibody-producing plasmablasts, with evidence of clonal cells in this cluster. Surprisingly, we did not detect plasmablast expansion in the blood of immunized individuals, despite a robust antibody response. This suggest that antibody-producing cells either migrate to their bone marrow niche at a time not captured by our weekly sampling, or remain in the tissues where they were generated. Recent studies have demonstrated that SARS-CoV-2 infection generates long-lived bone marrow plasma cells however, it remains to be elucidated whether mRNA vaccines drive a similar response (Nguyen et al., 2022; Turner et al., 2021a). Further studies evaluating the presence of SARS-CoV-2-specific, long-lived plasma cells in the bone marrow following immunization would shed light on the durability of protective immunity and aid in vaccine development.

Recent studies have demonstrated that SARS-CoV-2 infection and mRNA vaccines elicit potent antigen-specific GC responses (Lederer et al., 2020; Turner et al., 2021b). Consistent with the idea of long-lived plasma cell trafficking to the bone marrow, convalescent patients have reduced plasmablasts in circulation relative to those with acute illness, and the SHM footprint in the repertoire of the remaining cells was significantly diminished from what we had observed at the peak of the disease.

In our study, plasmablasts in COVID-19 patients were characterized by a strong IFN-I signature relative to those in healthy volunteers. While it has been shown that an overzealous IFN response favors extrafollicular plasma cell differentiation at the expense of affinity maturation during an anti-viral response (Braun et al., 2002; Soni et al., 2020), we did observe an accumulation of SHMs in the repertoire of plasmablasts and memory cells from COVID-19 patients, as well as in vaccinated individuals. We ought to consider that our observation of SHMs in plasmablasts may reflect the fact that we are studying patients with less severe disease; an earlier study of postmortem thoracic lymph nodes that described muted GC response was conducted in patients with severe COVID-19 (Kaneko et al., 2020). In our COVID-19 patients, clonal responses were most evident among plasmablasts. On the other hand, clonal cells were found within memory and resting B cells at multiple time points in vaccinated individuals. In future studies, stratifying patients by disease severity and a more detailed time course following vaccination should allow us to truly discern the impact of IFN on GC maturation in the context of SARS-CoV-2 infection and vaccination.

A number of studies have highlighted the shared IFN-induced gene signature in lymphocytes from patients with autoimmune disease and in subjects following viral infections (Kyogoku et al., 2013; Rönnblom and Leonard, 2019). Our observation that B lymphocyte transcriptional programs in COVID-19 patients are dominated by a marked upregulation of IFN-response genes may be important for understanding the immunopathology of COVID-19. Dysregulation of IFN-I signaling is a common factor in multiple autoimmune diseases, and there is growing evidence that autoantibodies could be driving severe disease and long-term sequelae in some COVID-19 patients (Acosta-Ampudia et al., 2022; Ehrenfeld et al., 2020; Psarras et al., 2017).

Based on the role of antigen-specific T cells in protective immunity against SARS-CoV-2 infection, it is becoming increasingly clear that successful vaccines need to engage the cellular adaptive immune response (Hirai and Yoshioka, 2022; Jeyanathan et al., 2020; Sauer and Harris, 2020). Indeed, humoral immune responses may be less effective against SARS-CoV-2 variants (Edara et al., 2021; Geers et al., 2021). Conversely, SARS-CoV-2-specific CD8 T cell responses, which target a broad range of epitopes, remain largely intact against variants (Liu et al., 2022; Moss, 2022). Our analysis revealed that both SARS-CoV-2 infection and, to a lesser degree, vaccination, elicit clonal CD8 effector T cell responses. We also observed a strong clonal response in CD8 T_EM_ cells in all volunteers following immunization – a feature of adaptive response that was notably absent in COVID-19 patients. This finding corroborates recent studies that have demonstrated that vaccination induces durable SARS-CoV-2-specific T cell responses (Hurme et al., 2022; Maringer et al., 2022; Sureshchandra et al., 2021).

Peripheral immune cells of COVID-19 patients were enriched in activated T cells, NK cells, and γδ T cells, with elevated expression of genes associated with cytotoxic effector functions (*GZMA*, *GZMB*, *GZMH*, *PRF1*, *GNLY*, *NKG7*, and *IL-32*). Clonal cytotoxic CD4 T cells in COVID-19 patients were largely absent in healthy volunteers following immunization. While hyperactivation of inflammatory responses and cytotoxic cells may contribute to immunopathology in severe illness, these features indicate protective immune responses and resolution of infection in mild and moderate disease (Chen and John Wherry, 2020). Furthermore, to our knowledge, our study is the first to highlight clonal expansion of γδ T cells in response to SARS-CoV-2 infection. Because few studies include analysis of γδ T cell repertoire, it remains to be elucidated whether these cells contribute to viral clearance or to the pathology associated with COVID-19.

This study underscores that SARS-CoV-2 infection and vaccination both lead to the development and maturation of antiviral adaptive immune responses. While the limited number of participants and absence of COVID-19 samples spanning a range of disease severities are important limitations of this study, we took steps to mitigate these limitations by validating our key findings with an analysis of a large publicly available dataset comprising 143 samples from healthy volunteers and COVID-19 patients. This dataset spans disease severity from asymptomatic to critical and supports the robustness of our findings (Stephenson et al., 2021). Future studies that include a granular analysis of a similarly high number of post-vaccination samples, preferably with time points immediately following the vaccine, will shed light on the differences in cytokine responses following these different types of immune challenges.

This study underscores the fine balance in COVID-19 between antiviral immune responses that achieve clearance of the infection and durable protective immunity, and those that lead to inflammation and immunopathology. Our highly granular data set enables direct comparison of immune responses in both infected individuals and those that received the mRNA SARS-CoV-2 vaccine and booster. Better understanding of the immunological features associated with protective immunity, immunopathology, and durability of protective immunological memory will aid not only in better viral-disease therapeutics, but also facilitate the development of effective vaccines for new and re-emerging viral diseases that threaten public health.

## Materials and methods

### Patients and sample collection

Peripheral blood samples were drawn from both outpatients and hospitalized patients with confirmed COVID-19 at NYU Langone Health. SARS-CoV-2 was detected in patients’ nasopharyngeal swab using the cobas^®^ SARS-CoV-2 real time PCR under EUA. Peripheral blood was collected in accordance with a NYU Institutional Review Board protocols (IRB 18-02035, 18-02037 and 20-00463). Samples were de-identified and assigned coded identification numbers prior to analysis.

Whole blood was collected in commercially available heparin-coated tubes (BD). Plasma was collected from whole blood by centrifugation at 2000 x *g* at 4°C, aliquoted, and stored at −20°C. For serum collection, whole blood was collected in serum separator tubes (SST). The blood was allowed to clot undisturbed at room temperature for 30-45 minutes and the clot was removed by centrifugation at 2000 x *g* at 4°C, the serum aliquoted and stored at −20°C.

### PBMC isolation

Peripheral blood mononuclear cells (PBMCs) were isolated from peripheral blood by diluting whole blood in gradient centrifugation using Ficoll-Paque PLUS (GE Healthcare) and SepMate™ PBMC Isolation Tubes (Stemcell) according to the manufacturer’s instructions. Buffy coat PBMCs were cryopreserved in FBS (Corning) supplemented with 10% DMSO (Sigma-Aldrich) and stored in liquid nitrogen.

### Single-cell RNA-seq

Sample processing of COVID-19 patient samples for scRNA-seq was performed in the ABSL3 facility of NYU Grossman School of Medicine (New York, NY), in accordance with its Biosafety Manual and Standard Operating Procedures. Single cell transcriptome profiling of PBMCs was carried out using the Chromium Next GEM Single Cell 5’ Library & Gel Bead Kit (v1.1 or v2) and Chromium controller (10x Genomics). PBMCs were thawed and rested for one hour at 4°C in RPMI 1640 with L-glutamine (Corning) supplemented with 20% fetal bovine serum (FBS). Live cells were enriched using Dead Cell Removal Kit (Miltenyi) to ensure viability of all samples is more than 95% prior to staining. To enable multiplexing and doublet detection, cells were stained with barcoded antibodies for CITE-seq and cell hashing described previously (Buus et al., 2020; Mimitou et al., 2019; Stoeckius et al., 2017; Stoeckius et al., 2018). Expression of selected surface protein markers (previously titrated antibody panel in supplemental materials) (Buus et al., 2020) was achieved by staining with barcoded antibodies as described (Stoeckius et al., 2017). Briefly, approximately 200,000 cells per sample were resuspended in staining buffer containing PBS (Corning), 2% BSA (Cytiva), 0.01% Tween (BioRad), and incubated for 10 minutes with Fc block (TruStain FcX, Biolegend; FcR blocking reagent, Miltenyi). Cells were then incubated with barcoded antibodies for 30 min at 4 °C. After staining, cells were washed four times in staining buffer. After the final wash, cells were resuspended in PBS + 0.04% BSA, filtered, and counted. Cells were pooled and loaded onto the Chromium chips (5 samples per lane, targeting 5,000 cells per sample). The single-cell capturing, barcoding, and cDNA library preparation were performed using the Chromium Next GEM Single Cell 5’ Library & Gel Bead Kit following the protocols recommended by the manufacturer. For experiments using Chromium Next GEM Single Cell 5’ Kit v1.1, HTO and ADT additive oligonucleotides were spiked into the cDNA amplification PCR and the ADT and HTO libraries were prepared as described previously (Stoeckius et al., 2017; Stoeckius et al., 2018). For experiments using Chromium Next GEM Single Cell 5’ Kit v2, the ADT library was prepared according to the manufacturer’s instructions. The cDNA fraction was processed according to the 10x Genomics Single Cell V(D)J protocol to generate the transcriptome library and the TCRα/β and BCR libraries. To amplify TCRγ/δ transcripts, we used a two-step PCR similar to the TCRα/β enrichment approach described previously (Mimitou et al., 2019). Libraries were pooled to desired quantities and sequenced on a NovaSeq 6000 100 cycle flow cell (R1: 26 cycles i5/i7: 10 cycles, R2: 74 cycles) (Illumina). Reads were trimmed as required for downstream processing.

### Single cell RNA-seq data processing

We used the Cell Ranger software suite from 10x Genomics (https://support.10xgenomics.com/single-cell-gene-expression/software/pipelines/latest/what-is-cell-ranger) to demultiplex cellular barcodes, align reads to the human genome (GRCh38 ensemble, http://useast.ensembl.org/Homo_sapiens/Info/Index), and perform UMI counting. We generated ADT and HTO count matrices using kallisto kb-count v0.24.1 (Bray et al., 2016; Melsted et al., 2021) and demultiplexed HTOs using HTODemux from Seurat v4.0.0 (Stuart et al., 2019). Following processing by Cell Ranger, all other processing was performed in R v4.0.3 (R Core Team, 2018). From filtered counts, Seurat was used to process the single-cell data, generate UMAP representation based on totalVI (Gayoso et al., 2021) dimension reduction of RNA and ADT modalities. RNA was normalized and batch-corrected by totalVI, while ADT values were corrected by the built-in integration function FindIntegrationAnchors in Seurat. Clustering was performed by the Louvain algorithm (Blondel et al., 2008) and cell type identification was determined by clustering, SingleR annotations, corrected ADT levels and canonical markers for various immune cell subsets. Further we performed sub-clustering on Myeloid, B, conventional T and innate-like T and NK cells to identify 32 individual populations. BCR/TCR sequences were processed by Cell Ranger VDJ and added to the metadata of the combined Seurat object for each sample. To evaluate the extent of somatic hypermutation in the B cell compartment, we used igBlast to identify mutations to the germline VH gene for each individual sample (https://www.ncbi.nlm.nih.gov/igblast/). Statistical differences in the percentages of cell type clusters were assessed by linear mixed models with disease and time point as fixed effects and subject as a random effect. To determine significance we used an ANOVA of each linear mixed model and post-hoc pairwise comparison of estimated marginal means. We performed gene set variation analysis (GSVA) of select immune cell subsets in R with the GSVA package v1.38 (Hänzelmann et al., 2013) with various named gene sets from MSigDB (http://www.gsea-msigdb.org/gsea/index.jsp) (Liberzon et al., 2015). We used the Wilcox test with a Benjamini-Hochberg p-value adjustment to assess differential expression among cell types and among cells from HCs, immunized individuals, and COVID-19 samples.

### Multiplex bead-binding assay for antibody profiling

Multiplex bead-binding assay was carried out as described (Hattori et al., 2021), with the following modifications. Briefly, we produced the spike and RBD proteins as described (Hattori et al., 2021) and purchased biotinylated nucleocapsid protein (Sino Biological, catalog number 40588-V27B-B). We used MultiCyt® QBeads® Streptavidin Coated panel QSAv1,2,3 and 5 (Sartorius catalog number 90792) to immobilize SARS-CoV-2 antigens; spike to QSAv1, Nucleocapsid to QSAv2, the receptor-binding-domain (RBD) of spike to QSAv3, and biotin only to QSAv5. The antigens were diluted to 25 nM in PBS with 0.5 % BSA and mixed with the same volume of the twice-washed QBeads. We detected antigen-specific antibodies in heat-inactivated serum or plasma using anti-human IgG-Alexa 488 (Jackson ImmunoResearch; catalog number 109-545-098, 1:800 in PBS 0.1 % Tween 20 and with 1 % BSA), anti-human IgA-PE (Jackson ImmunoResearch; catalog number 109-115-011, 1:100) and anti-human IgM-DyLight405 (Jackson ImmunoResearch; catalog number 709-475-073, 1:200). We measured the samples on a Yeti ZE5 Cell Analyzer (Bio-Rad) and analyzed the data using FlowJo (BD, version 10.7.1).

### ELISA

We used direct ELISA to quantify Spike-specific antibody titers in healthy volunteers before and after receiving BNT162b2 mRNA vaccine and booster as described (Samanovic et al., 2022). Briefly, 96-well plates were coated with 1 μg/ml S1 protein diluted in PBS and incubated overnight at 4°C (V08H and 40588-V08B, Sino Biological). Plates were washed with PBS containing 0.05% Tween-20 (PBS-T) and blocked with 5% non-fat milk in PBS-T. Sera were heat-inactivated at 56 °C for one hour, diluted in blocking solution, and added to the coated plate. After two hours’ incubation, the plate was washed with PBS-T. Horseradish peroxidase (HRP)-conjugated anti-human IgG antibody was diluted in blocking buffer, added to each well, incubated for one hour at room temperature, and washed with PBS-T. After developing with TMB Peroxidase Substrate 3,3′,5,5′-Tetramethylbenzidine (Thermo Fisher Scientific) for five minutes, we stopped the reaction with 1N hydrochloric acid and determined the optical density by measuring absorbance at 450 nm on a Synergy 4 (BioTek) plate reader.

### Validation Dataset

We used a public scRNA-seq dataset to validate some of the key findings of the study (Stephenson et al., 2021). The dataset includes 143 samples from healthy volunteers and COVID-19 patients, stratified by disease severity. In our analysis of data from Stephenson et al., we included COVID-19 samples with mild, moderate, severe, and critical disease.

## Data Availability

Data will be publicly available upon publication in peer reviewed journal.

## Acknowledgements

We are grateful for support of this work from NYU Grossman School of Medicine. Work in Dr. Koralov’s laboratory was further supported by the NIH (HL125816, CA271245, 2R44AI136141), LEO Foundation Grant (LF-OC-20-000351), NYU Cancer Center Pilot Grant (P30CA016087), the Judith and Stewart Colton Center for Autoimmunity Pilot grant. Presented work was also supported by NIH grant R21 AI158997, R01 CA194864 and R01 CA212608 to S.K.; NIH grants AI114852 and AI082630 to R.S.H.; K08AI163457 to R.J.U.; and AI148574 to M.J.M. TBB and NØ received support from the Danish Cancer Society (Kræftens Bekæmpelse), the Danish Council for Independent Research (Danmarks Frie Forskningsfond) and the LEO Foundation. We thank all members of NYU Vaccine Center processing and clinical staff, including Michael Tuen, Jimmy Wilson, Abdonnie Holder, Shelby Goins, Meron Tasissa, Sara Wesley Hyman, and Farzana Antara. We are also grateful to the NYU Genome Technology Core, and Dr. Heguy in particular, for technical assistance and support. We sincerely thank Ms. Cathy Shufro for her valuable contributions to improving the manuscript. Finally, we would like to thank all the study participants who have contributed to our studies.

## Disclosures

MJM reported potential competing interests: laboratory research and clinical trials contracts with Lilly, Pfizer (exclusive of the current work), and Sanofi for vaccines or MAB vs SARS-CoV-2; contract funding from USG/HHS/BARDA for research specimen characterization and repository; research grant funding from USG/HHS/NIH for SARS-CoV-2 vaccine and MAB clinical trials; personal fees from Meissa Vaccines, Inc. and Pfizer for Scientific Advisory Board service. RSH has received research support from CareDx for SARS-CoV-2 vaccine studies and has performed consulting work for Bristol-Myers-Squibb.

## Notes

### Clinical Trial

Observational trials are not subject to registration requirements

### Funding Statement

We are grateful for support of this work from NYU Grossman School of Medicine. Work in the Koralov laboratory was further supported by the NIH R01 grant (HL-125816), LEO
Foundation Grant (LF-OC-20-000351), NYU Cancer Center Pilot Grant (P30CA016087), the
Judith and Stewart Colton Center for Autoimmunity Pilot grant. Presented work was also
supported by NIH grant R21 AI158997, R01 CA194864 and R01 CA212608 to S.K.; NIH grants
AI114852 and AI082630 to R.S.H.; and AI148574 to M.J.M. TBB and NO are supported by the
Danish Cancer Society (Kraeftens Bekaempelse), the Danish Council for Independent Research (Danmarks Frie Forskningsfond) and the LEO Foundation.

### Author Declarations

NYU Institutional Review Board

### Summary of Updates

Additional samples were added to the analysis

