## Supplemental Table 1 for "mRNA COVID-19 vaccine elicits potent adaptive immune response without the persistent inflammation seen in SARS-CoV-2 infection"

| Participant | Cohort | Age | Sex | Race | BMI | Days post onset symptoms or days since first vaccine dose | Fraction inspired $O_2$ | Days since booster | WHO COVID severity scale | COVID Outcome | Co-morbidities |
|---|---|---|---|---|---|---|---|---|---|---|---|
| SK-010 | Acute COVID-19 | 36-40 | Female | Asian | NA | 12 | 21 | NA | 5 | Recovered | NA |
| | | | | | | 24 | 21 | | | | |
| SK-011 | Acute COVID-19 | 56-60 | Female | Caucasian | 35.7 | 6 | 32 | NA | 7 | Recovered | Type 2 Diabetes, Hypertension, Hyperlipidemia, Obesity |
| | | | | | | 13 | 24 | | | | |
| SK-012 | Acute COVID-19 | 41-45 | Male | Caucasian | 24.7 | 8 | 38 | NA | 8 | Recovered | Asthma, Anxiety, Depression |
| | | | | | | 24 | 38 | | | | |
| SK-013 | Acute COVID-19 | 61-65 | Male | NA | NA | 6 | 38 | NA | 5 | Recovered | NA |
| | | | | | | 11 | 28 | | | | |
| SK-014 | Acute COVID-19 | 51-55 | Female | Asian | NA | 9 | 21 | NA | 5 | Recovered | NA |
| CV-001 | mRNA vaccine | 36-40 | Male | Asian | 28.4 | 0, 7, 14,21, 28, 35 | 21 | 0, 7, 28 | NA | NA | NA |
| CV-003 | mRNA vaccine | 31-35 | Male | Asian | 27.9 | 0, 10, 20, 28, 35 | 21 | 0, 7, 28 | NA | NA | Hypothyroidism |
| CV-011 | mRNA vaccine | 36-40 | Male | Caucasian | 28.6 | 0, 7, 14, 21, 29, 36 | 21 | 0, 7 | NA | NA | NA |
| CV-012 | mRNA vaccine | 51-55 | Female | Caucasian | 28.7 | 0, 7, 21, 28 | 21 | NA | NA | NA | NA |
| CV-022 | mRNA vaccine | 41-45 | Male | Caucasian | 22.4 | 0, 7, 21, 28 | 21 | NA | NA | NA | Facial Cellulitis, GERD |
| CV-053 | mRNA vaccine | 46-50 | Female | African-American | NA | 0, 7, 21, 28 | 21 | NA | NA | NA | Fibroids, HSV, GERD |
| CV-056 | mRNA vaccine | 51-55 | Female | Caucasian | 25.4 | 0, 7, 21, 28 | 21 | NA | NA | NA | NA |
| SK-007 | HC | 36-40 | Female | Caucasian | 20.4 | 0 | 21 | NA | NA | NA | NA |
| SK-008 | HC | 36-40 | Female | Caucasian | 28.6 | 0 | 21 | NA | NA | NA | NA |

**Supplemental Table 1. Demographic characteristics, clinical features, and outcomes for study participants.**
