## Supplemental Figure 1 for "mRNA COVID-19 vaccine elicits potent adaptive immune response without the persistent inflammation seen in SARS-CoV-2 infection"

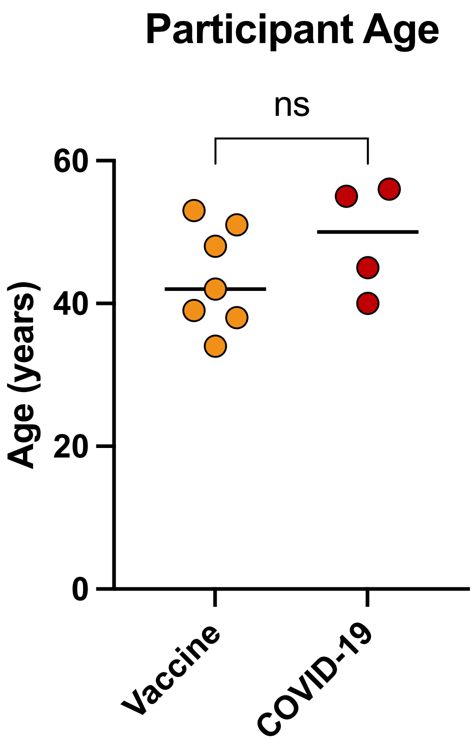


**Supplemental Figure 1. Participant age.**
Scatter plot of age for all healthy volunteers who received SARS-CoV2 vaccine and COVID-19 patients in **Supplemental Table 1**. P-value were determined by Welch's t-test (* $p < 0.05$, ** $p < 0.01$, *** $p < 0.001$).
