## Supplemental Figure 2 for "mRNA COVID-19 vaccine elicits potent adaptive immune response without the persistent inflammation seen in SARS-CoV-2 infection"

## A

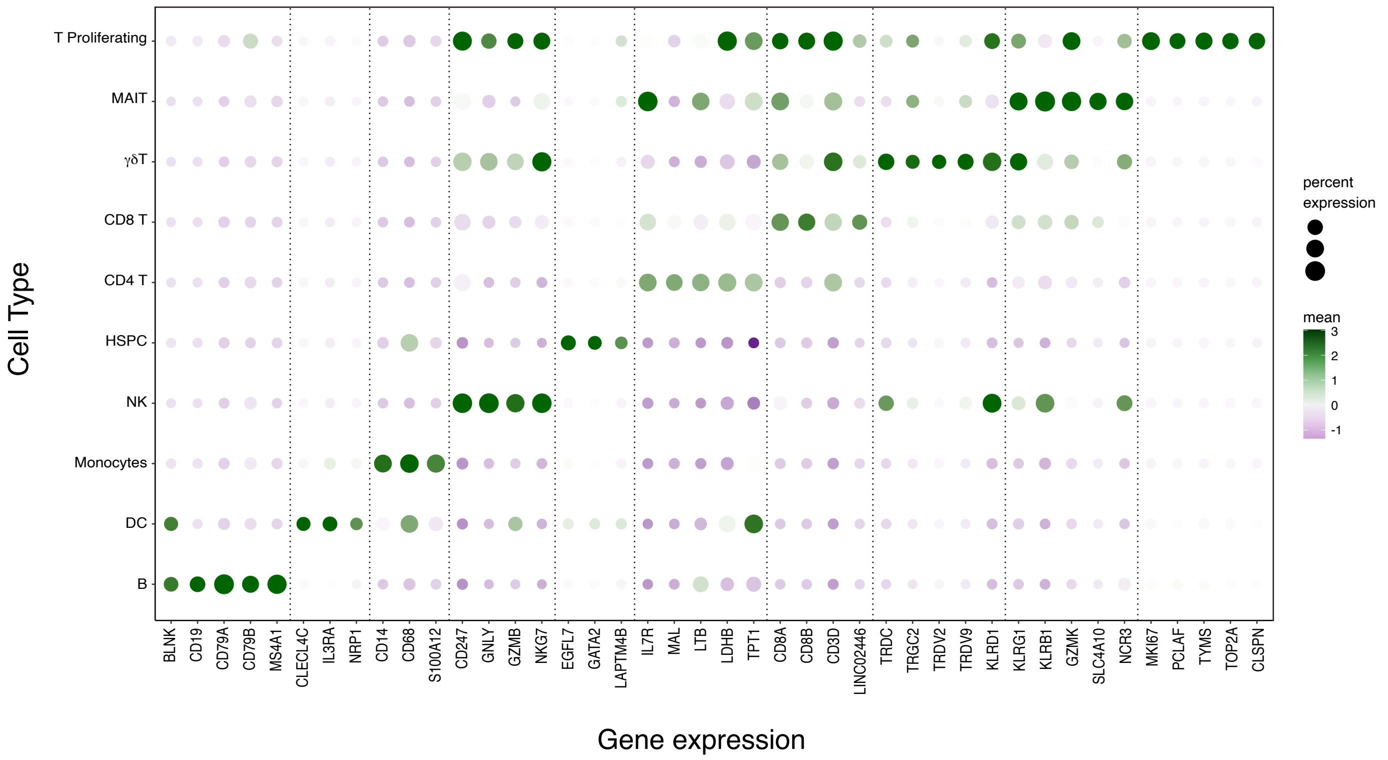


## B

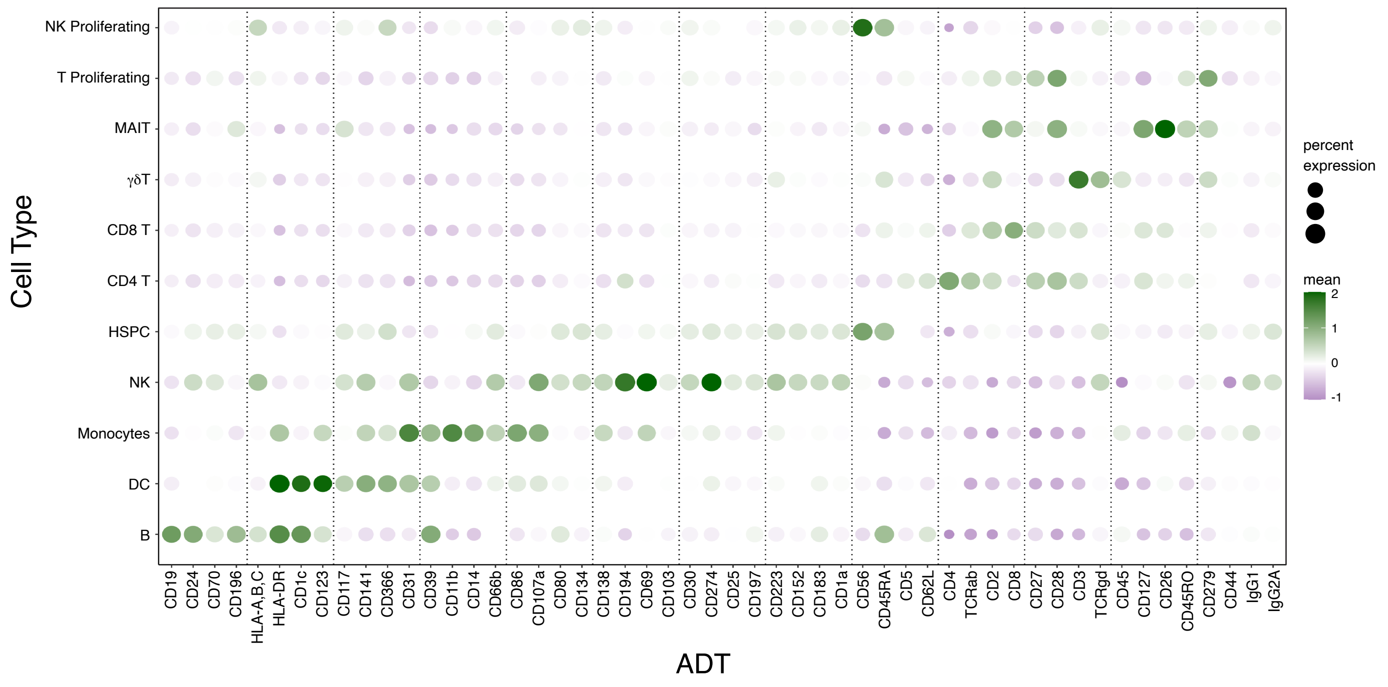

**Supplemental Figure 2. Surface protein and gene expression across identified populations.**
**A.** Dot plot illustrating the expression of key markers across the major cell populations identified. The gene names are shown on the x-axis and the identities of the cell populations on the y-axis. Downregulated markers are colored in purple and upregulated markers in green. The size of the dots represents the fraction of cells expressing the specific marker and the color intensity represents the average expression level.

**B.** Dot plot illustrating the distribution of antibody-derived tags (ADT) expression across the major cell populations identified. The protein markers are shown on the x-axis and the identities of the cell populations on the y-axis. Downregulated markers are colored in purple and upregulated markers in green. The size of the dots represents the fraction of cells expressing the specific marker and the color intensity represents the average expression level.
