## Supplemental Table 2 for "mRNA COVID-19 vaccine elicits potent adaptive immune response without the persistent inflammation seen in SARS-CoV-2 infection"

| Cell Population | Subpopulation | Markers |
| --- | --- | --- |
| B cells | Memory | MS4A1, COCH, AIM2, BANK1, CD79A |
| | Resting | IL4R, CXCR4, BTG1, TCL1A, YBX3 |
| | Plasmablasts | MZB1, TNFRSF17, CPNE5, POU2AF1 |
| | Activated | FOSB, CD196 (ADT) |
| T cells | T Proliferating | MKI67, TOP2A, PCLAF, TYMS, RRM2 |
| | CD4 T CM | CD4, TCF7, IL7R |
| | CD4 T Regulatory | CD4, CTLA4, FOXP3, IL2RA |
| | CD4 Activated | CD4, CCL5, FYB1, GZMK, IL32, TRAC, KLRB, GZMA, ZEB2 |
| | CD8 Naïve | CCR7, CD8A, CD8B |
| | CD8 EM | CD8A, CD28, CD127, CD45RO, CCL5, NKG7 |
| T innate cells | MAIT | TRAV1-2, SLC4A10, KLRB1, GZMK, IL7R, SLC4A10, CXCR6, CD45RO (ADT) |
| | NK | GNLY, TYROBP, NKG7, FGFBP2, KLRF1 |
| | γδT | TRDC, TRGC2, KLRC1, NKG7, TRDV2, TRGV9, KLRG1 |
| | NK Proliferating | GNLY, NKG7, MKI67, TOP2A, STMN1, TYMS |
| Myeloid cells | pDC | CLEC4C, IL3RA |
| | Classical Monocytes | CD14, LYZ, CD36 |
| | $NKG7^{+}$ Monocytes | LYZ, NKG7, GNLY |
| | Non-classical monocytes | FCGR3A, MS4A7, CDKN1C |
| | $NEAT1^{hi}$ Monocytes | CD14, NEAT1 |

**Supplemental table 2. Markers used to delineate immune cell subsets.**
