## Supplemental Figure 3 for "mRNA COVID-19 vaccine elicits potent adaptive immune response without the persistent inflammation seen in SARS-CoV-2 infection"

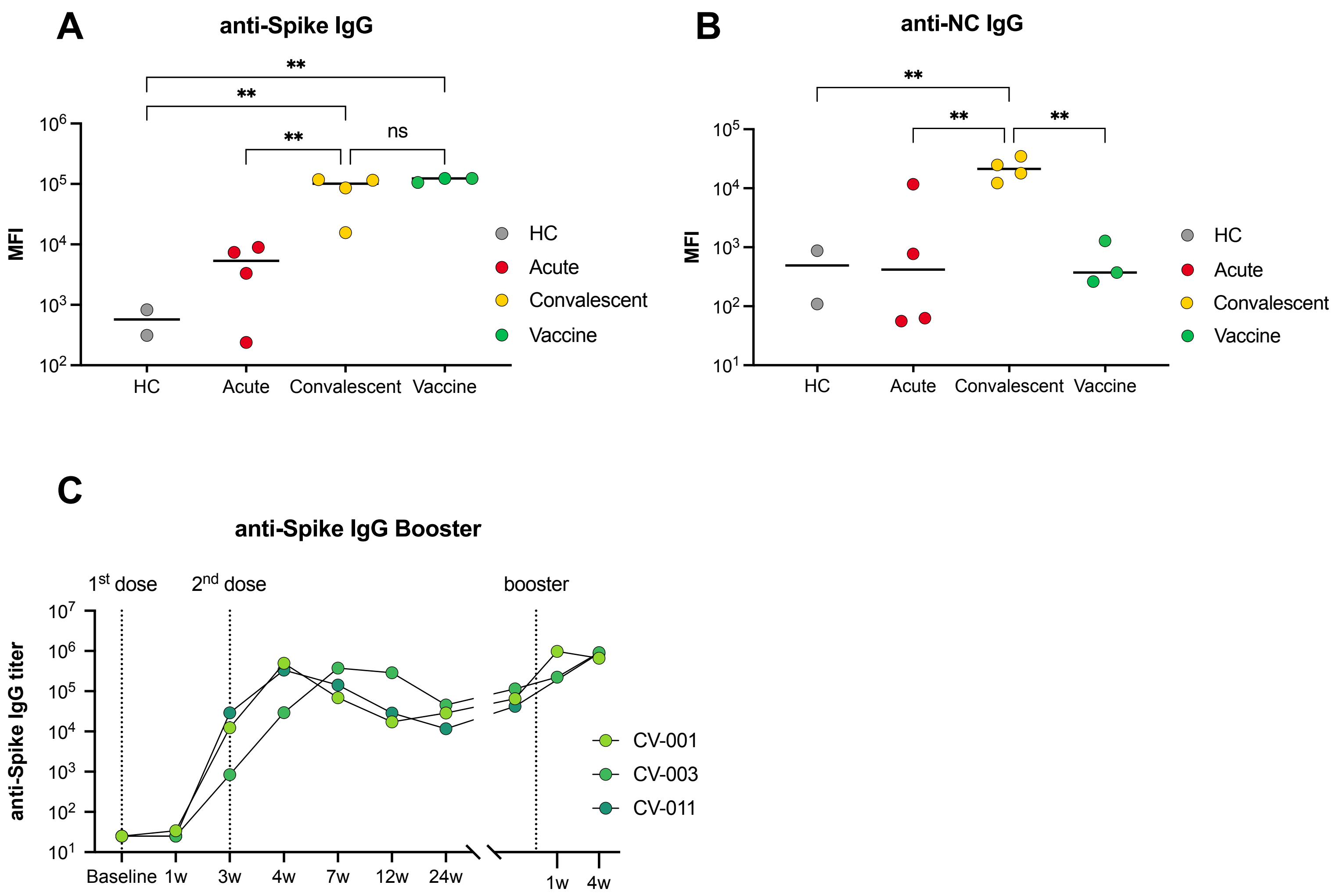


**Supplemental Figure 3. Ab titers and plasmablast responses**
**A-B.** SARS-CoV-2-specific Ab titers were assessed for COVID-19 patients and healthy volunteers using Multiplex Bead Binding Assay (MBBA). IgG anti-Spike responses are shown in (**A**), anti-NC responses in (**B**). COVID-19 patient samples are split by days post-onset (DPO) of symptoms into acute (≤10 DPO) and convalescent (>10 DPO). For the vaccine group, samples collected 4 weeks post-first vaccine dose were used. P-value were determined by Welch's t-test (* $p < 0.05$, ** $p < 0.01$, *** $p < 0.001$).

**C.** Spike-specific Ab titers for 3 healthy volunteers before and after receiving the BNT162b2 mRNA vaccine and booster, assessed by direct ELISA.
