## Supplemental Figure 4 for "mRNA COVID-19 vaccine elicits potent adaptive immune response without the persistent inflammation seen in SARS-CoV-2 infection"

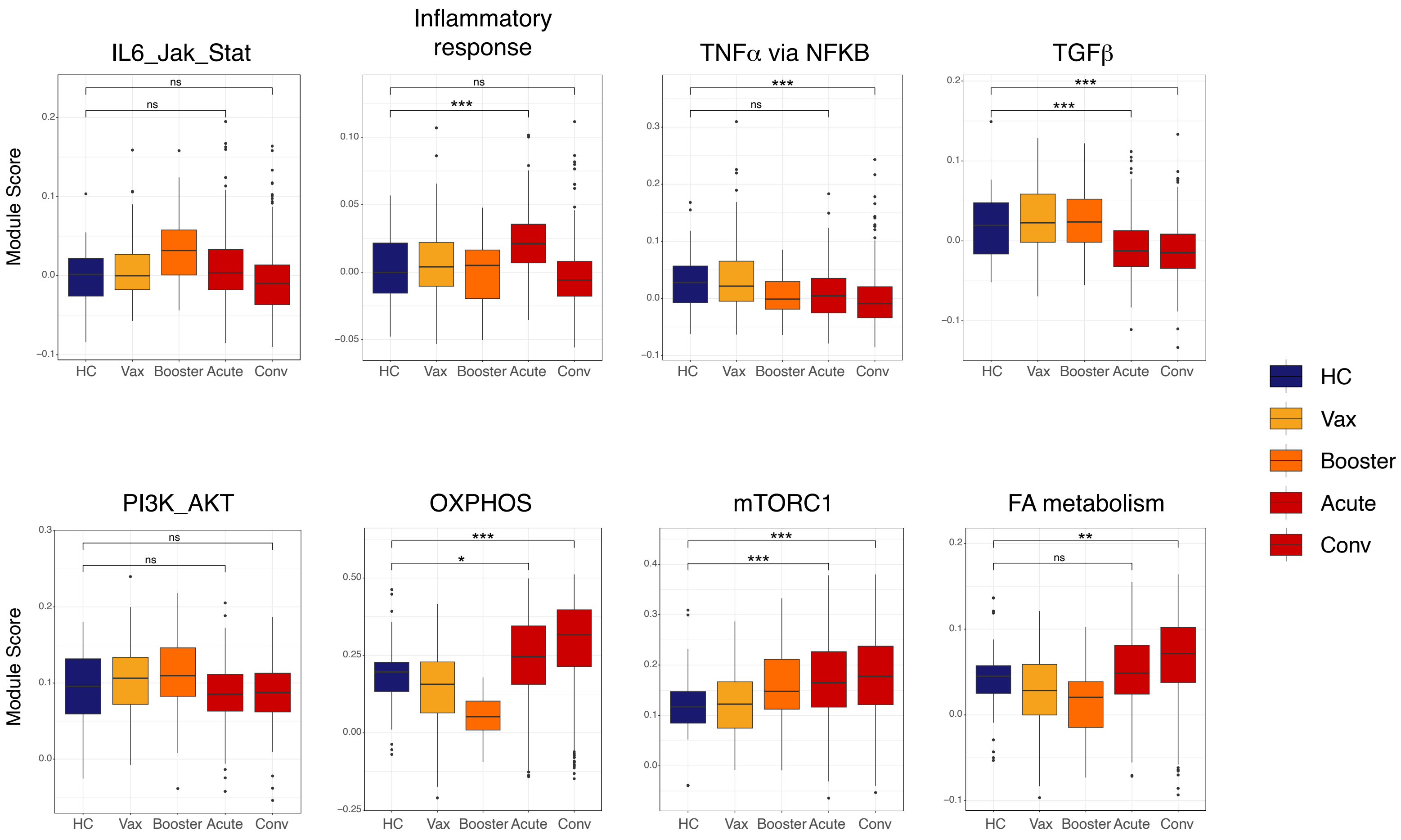


**Supplemental Figure 4. Pathway enrichment in plasmablasts module score.**
Module scores for the pathways in **Fig. 2A** were calculated using the AddModuleScore function from the Seurat package, which calculates the mean expression for a set of genes and adjusts for the collective expression of control features. The p-values were computed using the Wilcoxon test to compare the median module scores across a range of conditions (* $p < 0.05$, ** $p < 0.01$, *** $p < 0.001$).
