## Supplemental Figure 5 for "mRNA COVID-19 vaccine elicits potent adaptive immune response without the persistent inflammation seen in SARS-CoV-2 infection"

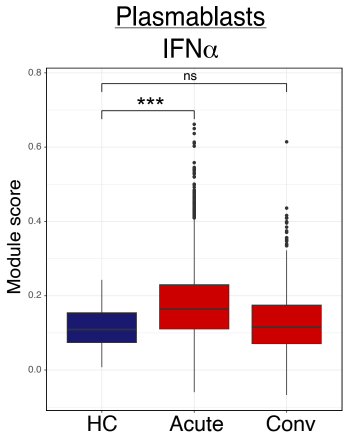


**Supplemental Figure 5. IFN-I gene pathway enrichment in plasmablasts module score in Haniffa data set.**
Module scores were calculated for plasmablasts in the Haniffa acute and convalescent COVID-19 samples (Stephenson*et al.*, 2021) using the AddModuleScore function from the Seurat package, which calculates the mean expression for a set of genes and adjusts for the collective expression of control features. The p-values were computed using the Wilcoxon test to compare the median module scores across a range of conditions (* $p < 0.05$, ** $p < 0.01$, *** $p < 0.001$).
