## Supplemental Figure 6 for "mRNA COVID-19 vaccine elicits potent adaptive immune response without the persistent inflammation seen in SARS-CoV-2 infection"

A

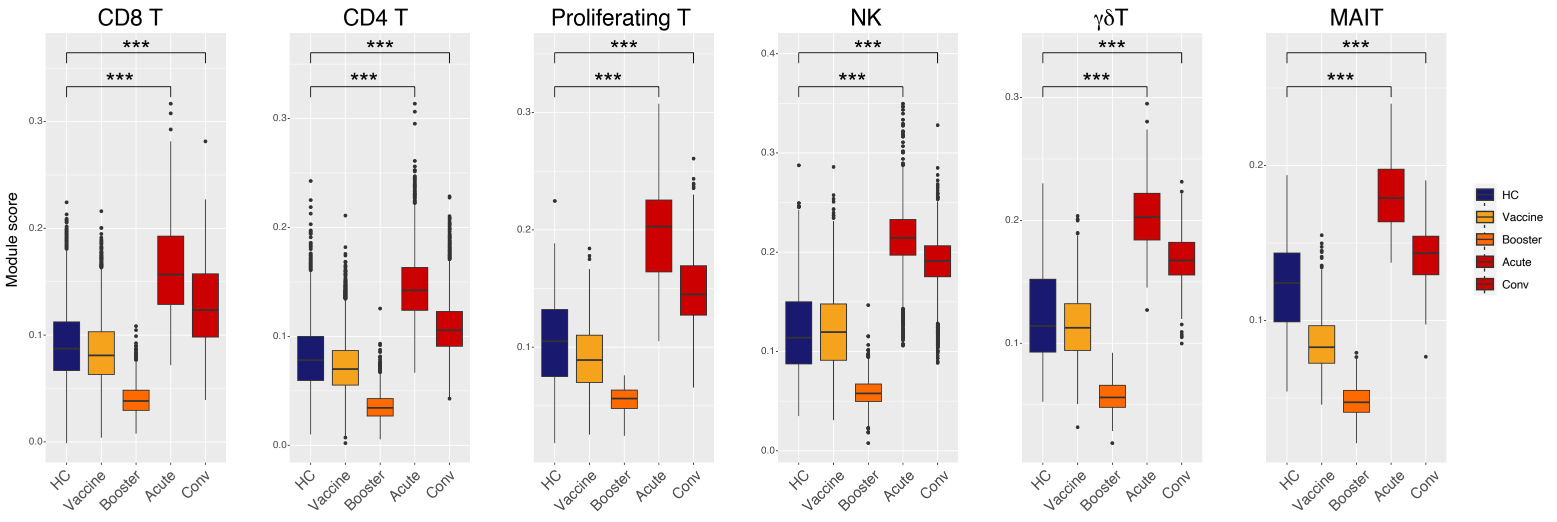


B

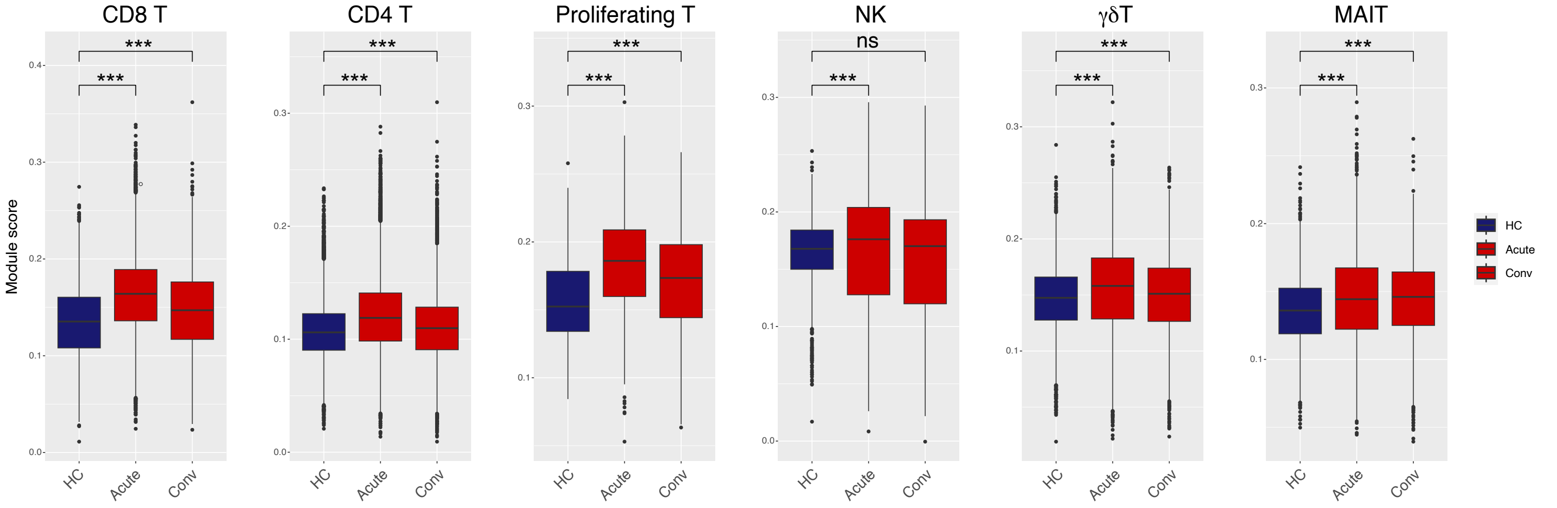


C

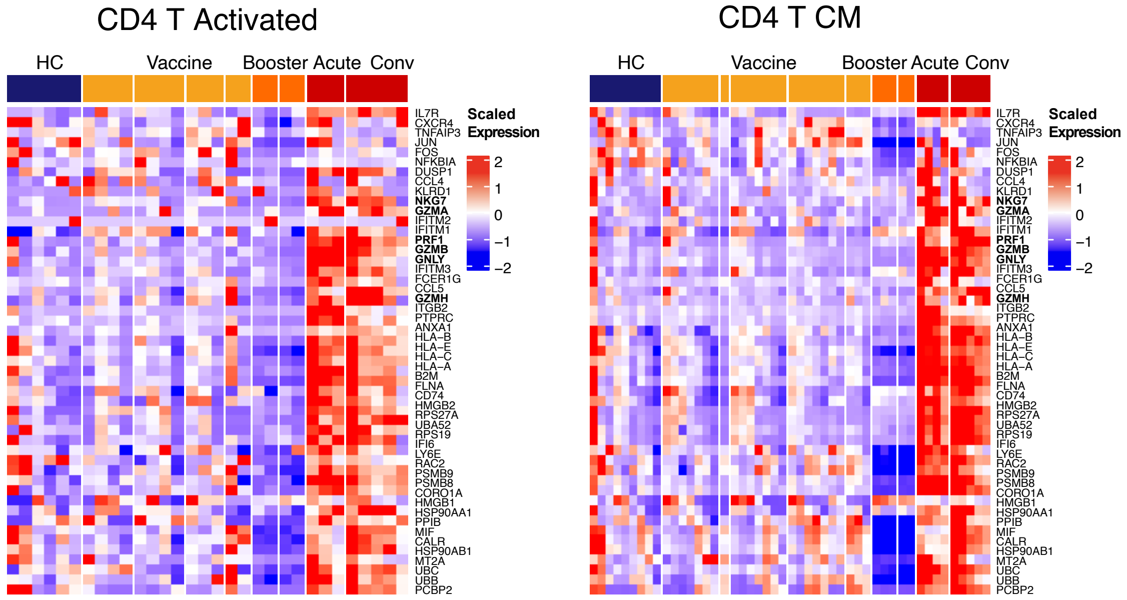

**Supplemental Figure 6. Cytotoxic responses and clonality of conventional and innate-like T cells in COVID-19 and SARS-CoV-2 vaccine recipients.**

**A,B.** Module scores were calculated based on expression of genes associated with cytotoxic effector function from the gene set T cell mediated cytotoxicity (GO:0001913) in major conventional and innate-like T cell populations in our data set (**A**) andvalidated in Haniffa data set (**B**)(Stephenson *et al.*, 2021)

**C.** Average per-sample scaled expression of genes associated with cytotoxic effector function from the gene set T cell mediated cytotoxicity (GO:0001913) in activated CD4 and CD4 CM T cells.
