## Supplemental Figure 7 for "mRNA COVID-19 vaccine elicits potent adaptive immune response without the persistent inflammation seen in SARS-CoV-2 infection"

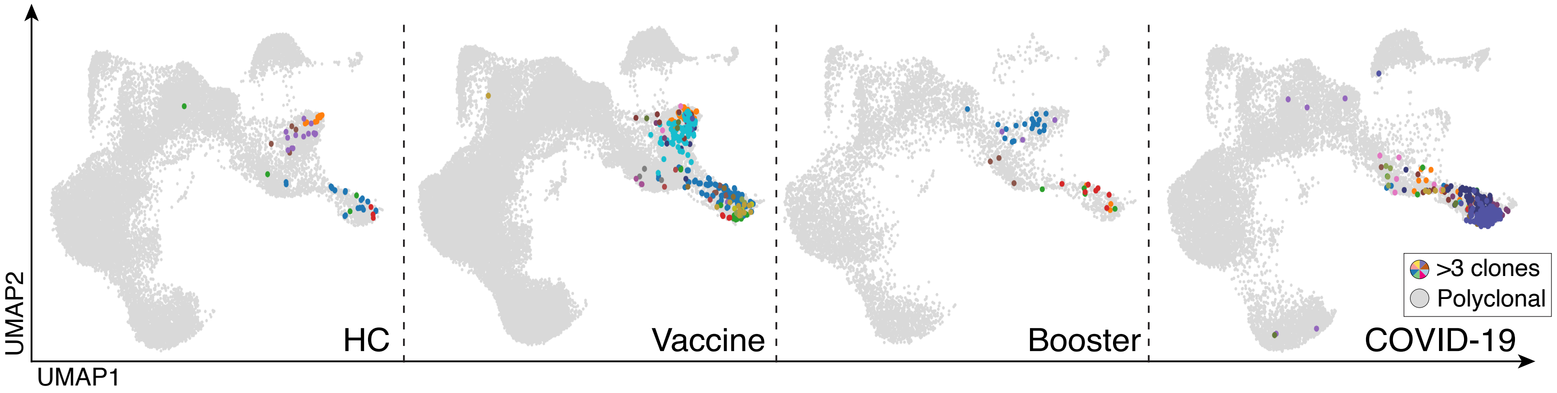


**Supplemental Figure 7. SARS-CoV-2 reactive CD8 T cell clones.**
UMAP visualization of CD8 T cell clones that match reported TCRβ sequences from natural and synthetic exposure to SARS-CoV-2. Since clusters CD8 T Effector Memory and CD4 T Activated overlap in UMAP space, we confirmed that all clonal cells were from the CD8 T Effector Memory population. At a 100% sequence identity threshold, clonal TCRab CDR3 amino acid sequences were compared to CDR3 sequences from spike-specific T cell clones using the cd-hit-2d command from the CD-HIT package. Identical CDR3 sequences in at least 3 cells that had matches to CDR3s in the AIM assay are colored uniquely.
