## Supplemental Figure 8 for "mRNA COVID-19 vaccine elicits potent adaptive immune response without the persistent inflammation seen in SARS-CoV-2 infection"

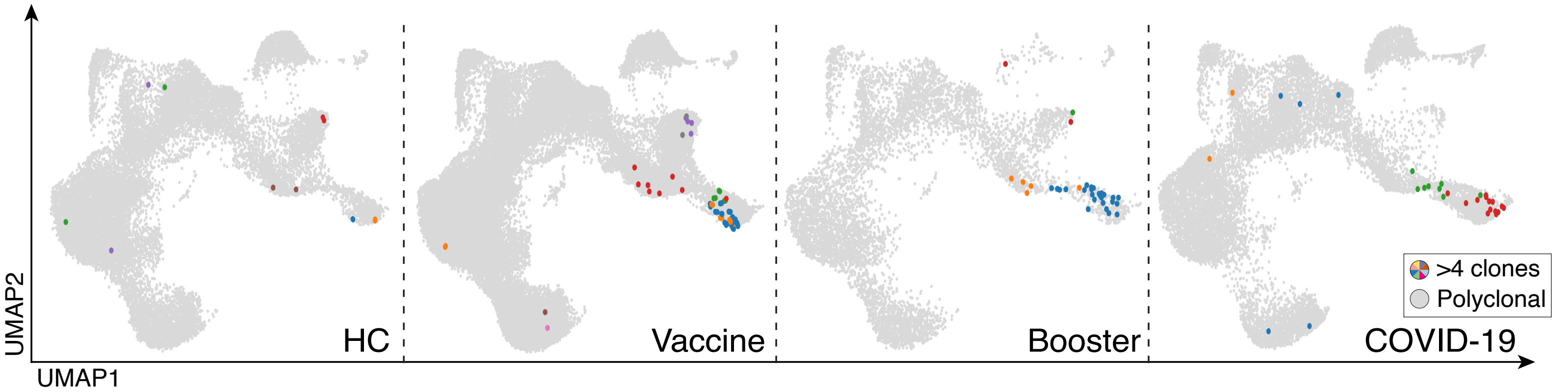


**Supplemental Figure 7. SARS-CoV-2 reactive CD4 T cell clones.**
UMAP visualization of T cells expressing TCRs with CDR3 sequences found in SARS-CoV-2 reactive T cells in antigen-induced activation (AIM) assay using Spike peptides. At a 95% sequence identity threshold, clonal TCRab CDR3 amino acid sequences were compared to CDR3 sequences from spike-specific T cell clones using the cd-hit-2d command from the CD-HIT package. Identical CDR3 sequences in at least 4 cells that had matches to CDR3s in the AIM assay are colored uniquely.
