## Supplemental Figure 9 for "mRNA COVID-19 vaccine elicits potent adaptive immune response without the persistent inflammation seen in SARS-CoV-2 infection"

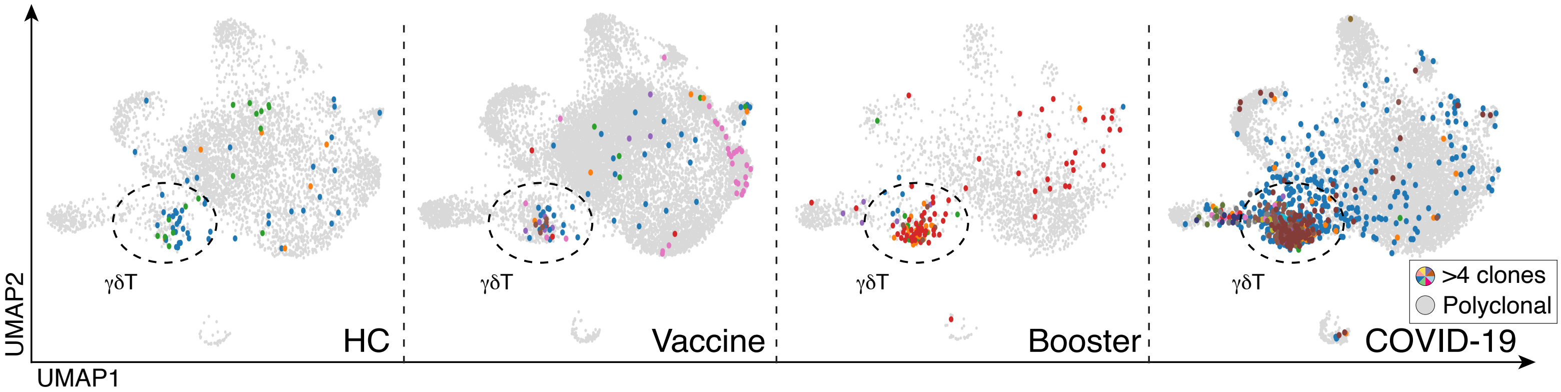


**Supplemental Figure 9. Clonality of γδT cell.**
UMAP visualization of clonal γδT cells from healthy volunteers before (first panel) and after (second panel) receiving the BNT162b2 mRNA vaccine and booster (third panel), and COVID-19 patients (fourth panel).
Clonality is determined by the CDR3 sequence in TCRδ chain. Identical CDR3 sequences in at least 5 cells are colored uniquely.
